# Adjusting confirmed COVID-19 case counts for testing volume

**DOI:** 10.1101/2020.06.26.20141135

**Authors:** Nathan Favero

## Abstract

When assessing the relative prevalence of the novel coronavirus (COVID-19), observers often point to the number of COVID-19 cases that have been confirmed through viral testing. However, comparisons based on confirmed case counts alone can be misleading since a higher case count may reflect either a higher disease prevalence or a better rate of disease detection. Using weekly records of viral test results for each state in the US, I demonstrate how confirmed case counts can be adjusted based on the percentage of COVID-19 tests that come back positive. A regression analysis indicates that case counts track better with future hospitalizations and deaths when employing this simple adjustment for testing coverage. Viral testing results can be used as a leading indicator of COVID-19 prevalence, but data reporting standards should be improved, and care should be taken to account for testing coverage when comparing confirmed case counts.

---

Policy makers and public health officials face numerous tradeoffs as they respond to the COVID-19 pandemic, including decisions about where to deploy certain medical or disaster relief resources, when and how severely to limit everyday activities that risk contributing to the spread of the disease, and what types of economic relief should be offered and for how long. Ordinary citizens also face personal decisions about what kinds of precautions they will take to limit the risk of being exposed to or of spreading the disease. One consideration for people weighing alternative courses of action amid the COVID-19 pandemic is the current prevalence of the disease in different geographic areas.^1^ Unfortunately, it is difficult to come by accurate real-time information about the current prevalence of active disease cases, in part because of unavoidable lags associated with most ways of measuring disease prevalence. Counts of COVID-19 deaths are widely cited, but these are known to be a lagging indicator of disease prevalence because in fatal cases of the disease there is often a delay of several weeks between initial infection and death.^2^ Data on the volume of hospitalizations should exhibit a smaller lag than data on deaths, but it can still lag behind initial detection of disease cases because among those eventually hospitalized, hospital admission often occurs several days after symptom onset.^3,4^ The availability of data on hospitalizations in the US also varies widely from state to state. Data on the number of new COVID-19 cases confirmed through viral (SARS-CoV-2) testing constitutes another source of information about disease prevalence. Confirmed case counts are reported more consistently across states than hospitalizations, although discrepancies in reporting standards have still been identified. For example, in mid-May, several states and the US Centers for Disease Control and Prevention (C<u>DC</u>) were found to be including antibody test results in their reports of confirmed cases.^5^ Unlike viral tests, antibody tests are not designed to test for current infection, so failing to distinguish between viral and antibody tests in their reporting led to misleading numbers being reported by several states—a problem which most states appear to have now remedied.^6^ Confirmed case counts are also subject to lags because of the delay between initial infection and when someone gets tested as well as the time it takes for test specimens to be analyzed and results reported. Nonetheless, case counts confirmed through viral testing probably constitute the COVID-19 prevalence indicator with the shortest lag time, among widely-reported indicators.

Despite the importance of confirmed case counts as a relatively early indicator of disease prevalence, under-detection of cases is a serious concern for this type of data since an increase in the reported number of confirmed cases may reflect an improvement in detection rates rather than an increase in the underlying prevalence of the disease.^7^ Indeed, the steady increase in the volume of viral tests being conducted in the US throughout the past several months has complicated the interpretation of trendlines in naïve case counts over time.^8^ Differences in testing volume also complicate comparisons of geographic units (e.g., countries, states, or counties) to one another since a higher volume of confirmed cases in one unit could merely reflect a better rate of detection.

Discussions of testing coverage sometimes point to data on the percentage of viral tests that have yielded positive reports. This simple indicator of testing coverage has been highlighted in several places, including the official CDC guidelines for reopening.^9^ While a low percentage of positive cases does not imply that all instances of the disease are being detected, it does suggest that testing coverage is broad enough to accommodate testing of not only those who are at greatest risk of having contracted the disease but also many people with relatively low odds of having contracted COVID-19. The percentage of positive tests is sometimes reported alongside the number of confirmed cases, which can aid interpretation of likely trends in disease prevalence.^10^ For example, if one observes decreases in both the number of confirmed cases and the percent of positive tests, this suggests that disease prevalence has decreased. At the same time, I am aware of no guidance from officials or the academic literature regarding how to rectify conflicting trendlines from the two indicators. If a state sees the number of newly-confirmed cases per week increase from 400 to 600 but the percent positive rate simultaneously decreases from 10% to 5%, is that a sign that prevalence has increased, decreased, or remained approximately steady?

The current study introduces a simple method for adjusting confirmed case counts based on testing coverage, which should allow for better comparisons of disease prevalence across time periods or across units when there is variance in testing coverage. Under this method, newly confirmed case counts are multiplied by a case multiplier (m), which is calculated as a linear function of the percentage of new viral tests that yield positive results (p):

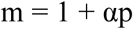

The value of the parameter α can be calibrated using a regression model in which the dependent variable is some other indicator of disease prevalence (e.g., future deaths or hospitalizations).

## Study Data and Methods

### Study Data

A state-level analysis was conducted to determine how well COVID-19 test results have tracked with future counts of COVID-19-related deaths and hospitalizations between March 8 and June 13, 2020. Appendix Table 1 shows descriptive statistics for all variable used in this analysis. The COVID Tracking Project maintains a publicly available dataset of daily state reports of COVID-19 test results.^11^ Weekly measures of the number of newly confirmed COVID-19 cases as well as the percentage of newly reported tests that are positive were constructed based on changes in the cumulative number of positive and negative tests reported on the last day of each week (or the latest day of the week for which non-missing values were available). The COVID Tracking Project primarily tracks the results of viral tests, although available reports have at times not distinguished between antibody and viral test results, meaning that a limited number of antibody test results are included among their counts for certain states and in certain weeks.^12^ Antibody tests are not designed to test for current infection, so the inclusion of a limited number of antibody test results in the dataset likely makes the COVID Tracking Project testing data a somewhat worse indicator of contemporaneous disease severity than it would be if cleaner testing data had been consistently available.

**Table 1.** Ordinary least squares regression results.

|  | COVID-19 deaths<sup>a,b</sup> |  | Excess select respiratory illness deaths<sup>a,b</sup> |  | Excess all-cause deaths<sup>a</sup> |  | Current COVID-19 hospitalizations<sup>c</sup> |  |
| --- | --- | --- | --- | --- | --- | --- | --- | --- |
|  | b/se | b/se | b/se | b/se | b/se | b/se | b/se | b/se |
| New cases | 0.065***<br>(0.0050) | 0.042***<br>(0.0047) | 0.066***<br>(0.0056) | 0.040***<br>(0.0051) | 0.093***<br>(0.0063) | 0.069***<br>(0.0089) | 0.25***<br>(0.015) | 0.20***<br>(0.024) |
| New cases ×<br>% Pos. |  | 0.00089***<br>(0.00017) |  | 0.0010***<br>(0.00017) |  | 0.00094**<br>(0.00031) |  | 0.0019**<br>(0.00054) |
| R-sqr | 0.867 | 0.905 | 0.847 | 0.892 | 0.766 | 0.784 | 0.894 | 0.907 |
| Implied case<br>multiplier<br>(m)<sup>d</sup> |  | 1 + 0.021p |  | 1 + 0.025p |  | 1 + 0.014p |  | 1 + 0.009p |
| N | 503 | 503 | 503 | 503 | 503 | 503 | 390 | 390 |

Notes: All counts measured per 100,000 population. Standard errors (se), in parentheses, are robust to clustering by state. Regressions were estimated without the inclusion of a constant term, and both independent variables were lagged 1 week. <sup>a</sup>Estimates of final death counts (the outcome) were created based on provisional death counts adjusted for state-specific reporting delays. The sample consists of weekly data from March 15-May 30, 2020 and includes 48 states plus the District of Columbia. Connecticut and North Carolina were excluded due to data irregularities. <sup>b</sup>Age-standardized through indirect standardization, using 2018 population estimates from the US Census Bureau and infection fatality rate estimates from Verity et al. (https://doi.org/10.1016/S1473-3099(20)30243-7). <sup>c</sup>The sample consists of weekly data from March 28-June 13, 2020 and includes 44 states plus the District of Columbia. Alabama, Florida,
Hawaii, Idaho, Kansas, and Tennessee did not report current COVID-19 hospitalizations and are not included in the sample. <sup>d</sup>See appendix for further details. \* $p<0.05$ , \*\* $p<0.01$ , \*\*\* $p<0.001$ (two-tailed).

### Outcomes of Interest

To assess the validity of using aggregated viral testing data as an indicator of COVID-19 prevalence, testing data was compared to four different outcome measures that should reflect COVID-19 outbreak severity. Three of the outcomes are measures of deaths likely attributable to COVID-19, and the fourth outcome is COVID-19 hospitalizations. Since these outcome measures are all expected to lag behind initial confirmation of COVID-19 cases, each week’s testing results are compared to deaths and hospitalizations recorded one week later.

The National Center for Health Statistics (NCHS) reports preliminary counts of weekly deaths based on death certificate data.^13^ Death certificates provide the most reliable source of mortality data and allow for deaths to be coded according to the date when a death actually occurred—not the date on which the death was reported as a COVID-19 death by state authorities. However, death certificate data can be subject to substantial reporting delays, resulting in provisional death counts that initially understate the number of deaths while processing of death certificate data is still ongoing.^14^ Fortunately, the vast majority of deaths in most states appear to be reported within the first few weeks after a death occurs, and estimates of final death counts can be made based on examining prior revisions to provisional counts. Estimates of expected revisions due to reporting delays were created by calculating average proportional increases in weekly all-cause death counts for each state based on the number of weeks between the report date and the reference week. Friday updates to provisional counts published from May 15 to June 19, 2020 were used to calculate average increases. Counts were only adjusted to account for delays observed in the first 16 weeks after the reference week, after which only small revisions to death counts were observed and data availability was more limited (since the COVID-19 death count file only includes deaths occurring since January 26, 2020). For the main analysis of how testing data is associated with death counts, deaths occurring after May 30 are not considered because of higher unreliability of provisional counts in recent weeks given shorter reporting times. NCHS suppressed death counts to protect privacy when between one and nine deaths were reported; these missing values were replaced with fives for this analysis. Connecticut and North Carolina were excluded from analyses using death counts due to data irregularities.

Given that deaths caused by COVID-19 may be misattributed to other causes, three different measures of death counts are used: a count of deaths attributed to COVID-19; an estimate of excess deaths attributed to select respiratory illnesses; and an estimate of excess all-cause deaths. The count of deaths identified as involving COVID-19 based on death certificate data was age-standardized through indirect standardization, using 2018 population estimates from the US Census Bureau and a recent estimate of infection fatality rate estimates by age.^15^ COVID-19 deaths may be especially likely to be misclassified as deaths caused by pneumonia or influenza because of overlapping symptoms,^13^ so an estimate of excess select respiratory illness deaths was also created by comparing the number of deaths attributed to COVID-19, pneumonia, and/or influenza (ICD-10 codes U07.1 or J09-18) to historical data on pneumonia and influenza deaths.^16^ A measure of all-cause deaths was also created, but this measure likely reflects some deaths that are indirectly attributable to the COVID-19 pandemic (e.g., because people avoiding the emergency room due to concerns about exposure to SARS-CoV-2, the virus that causes COVID-19).^17,18,19^ Both excess death measures were calculated as the number of observed deaths minus the average number of deaths during the same week in the prior three years (2017-2019). The select respiratory cause excess deaths measure was age-standardized using the same method as for COVID-19 death counts, but the all-cause excess deaths measure was not age-standardized because the measure partially reflects deaths indirectly attributable to COVID-19.

Data on the number of current COVID-19 hospitalizations was obtained from the COVID Tracking Project, which started reporting this variable during the week of March 28, 2020.^11^ This hospitalization variable was available for 44 states plus the District of Columbia; many of these states have missing values for earlier weeks of the sample because they were not yet reporting this information. Since the COVID Tracking Project publishes daily data, a weekly state-level dataset was constructed using the last day of the week for which a non-missing value of hospitalizations was available.

### Analysis

All count variables were transformed to indicate prevalence as a rate, taking the form of a count per 100,000 people. Ordinary least squares regression was used to estimate a separate model for each death or hospitalization outcome, with data from the prior week’s reported COVID-19 test results being used to predict the current week’s outcome. All analysis was conducted using Stata SE (version 16.1). Full replication materials can be found at https://github.com/favero-nate/covid-underreporting.

### Limitations

Perhaps the most notable limitations of this study relate to the known sources of measurement error present in each of the variables being analyzed. There are known inconsistencies in reporting standards for COVID-19 testing and for outcome measures across states, so the relationship between test results and outcomes may differ somewhat from one state to another. By pooling all states together in a single analysis, this research can illuminate the general nationwide tendency for testing results to track with outcome measures, but it will not illuminate the nature of any idiosyncrasies distinct to particular states. The death counts used in the analysis are based on provisional data, making it necessary to make adjustments for reporting delays. These adjustments will not be perfect and constitute another source of error. For example, if a state reports deaths for one particular week more quickly than is typical, this may result in an overestimation of the true number of deaths for that week. This source of error will disproportionately affect the most recent weeks included in the analysis since less adjustment for reporting delays is necessary after more time has passed (allowing greater opportunity for missing deaths to be reported). There is also noisiness in the estimated death estimates caused by year-to-year variance in the volume of deaths due to factors other than COVID-19. One additional source of noise is the fact that available data sources do not allow for precise adjustments for the extent to which the link between disease prevalence and the outcomes observed here depends on the age or relevant comorbidities of those contracting COVID-19 within a given state. To the extent that indicators are noisy, we might expect them to generally weaken the precision with which relationships can be estimated and to push the results toward null findings, although other biases in estimates can occur when measurement errors are correlated with one another. Since the outcome measures are generally detected and reported through processes distinct from those of the predictors (especially in the case of all-cause excess deaths), correlated measurement error is not expected to be a serious problem. It is also worth noting that this analysis is retrospective and does not test the ability of a model using viral test results as predictors to make a true forecast of future outcomes.

## Study Results

Table 1 displays full regression results for the analysis, which indicate that confirmed COVID-19 case counts track fairly well with death and hospitalization outcomes in the following week. Furthermore, the exact relationship between confirmed case counts and outcomes is dependent on the percentage of tests that yield positive results—an indicator of testing coverage. Specifically, the explanatory power of the regression model improves noticeably (as reflected in the models’ R-squared values) whenever an adjustment for the percent of tests that are positive is included in the regression, and the coefficient for this adjustment is statistically significant (at the .01 level) in each regression.

The extent to which the relationship between cases counts and outcomes depends on testing coverage is demonstrated in Table 2, which provides estimated quantities of each outcome per 100 confirmed COVID-19 cases. Whenever a higher percentage of viral tests return positive results, 100 confirmed cases of COVID-19 is associated with a higher number of expected deaths or hospitalizations one week later. For example, when the 5% of tests are positive, every 100 confirmed COVID-19 cases is associated with an estimated 4.61 COVID-19 deaths in the following week. But when 45% of tests are positive, 100 confirmed cases is associated with 8.19 COVID-19 deaths one week later. Compared to when the positive rate for COVID-19 tests is 5%, a 45% positive test rate is associated with between 33% and 81% more deaths or hospitalizations per 100 confirmed COVID-19 cases, depending on which of the four outcomes is being considered.

**Table 2.** Expected number of deaths or hospitalizations per 100 newly confirmed COVID-19 cases in prior week (95% confidence intervals)

|  | COVID-19<br>deaths<sup>a</sup> | Excess select<br>respiratory<br>illness deaths<sup>a</sup> | Excess all-cause<br>deaths<sup>a</sup> | Current COVID-19<br>hospitalizations<sup>b</sup> |
| --- | --- | --- | --- | --- |
| **At positive**<br>**test rate:** |  |  |  |  |
| 5% positive | 4.61 (3.80-5.43) | 4.55 (3.66-5.45) | 7.38 (5.84-8.91) | 21.4 (17.2-25.6) |
| 15% positive | 5.51 (4.87-6.15) | 5.56 (4.87-6.26) | 8.32 (7.23-9.41) | 23.3 (20.1-26.4) |
| 25% positive | 6.40 (5.80-7.01) | 6.58 (5.93-7.22) | 9.26 (8.37-10.1) | 25.1 (23.0-27.3) |
| 35% positive | 7.29 (6.55-8.04) | 7.59 (6.81-8.36) | 10.2 (9.14-11.3) | 27.0 (25.8-28.2) |
| 45% positive | 8.19 (7.21-9.16) | 8.60 (7.59-9.61) | 11.1 (9.65-12.6) | 28.9 (28.0-29.8) |

Notes: Estimates derived from regression models shown in table 1. <sup>a</sup>n=503. <sup>b</sup>n=390.

The effect that adjusting for the positive test rate has on the precision with which confirmed case counts track with future outcomes can be illustrated visually by examining a plot of nationwide counts new COVID-19 deaths over time (Figure 1). Predicted values of COVID-19 deaths based on the naïve confirmed case count are compared to predictions based a version of the confirmed case count that is adjusted for the percentage of positive tests (based on the previous state-level regression results). Case counts from the prior week are used to predict deaths, so the data in the exhibit has been adjusted to account for the lag between reports of new cases and the occurrence of COVID-19 deaths. Relying on the naïve case count leads to an underestimation of the number of COVID-19 deaths throughout April, during which time the testing coverage was poor and the percentage of positive tests mostly hovered at or near 20%. During the month of May, the testing coverage improved dramatically, with the percentage of positive tests reaching 6% by May 23. The increasing testing coverage throughout May means that a greater share of cases were likely being detected. During this expansion of testing coverage, the naïve number of newly confirmed cases appears to decrease very slightly over time, almost resembling a plateau. However, the number of COVID-19 deaths fell noticeably throughout the month of May, suggesting that prevalence of COVID-19 was actually declining substantially during this time. This decline in deaths is more closely (but still imperfectly) approximated by the case count that was adjusted to account for testing coverage.

**Figure 1.**
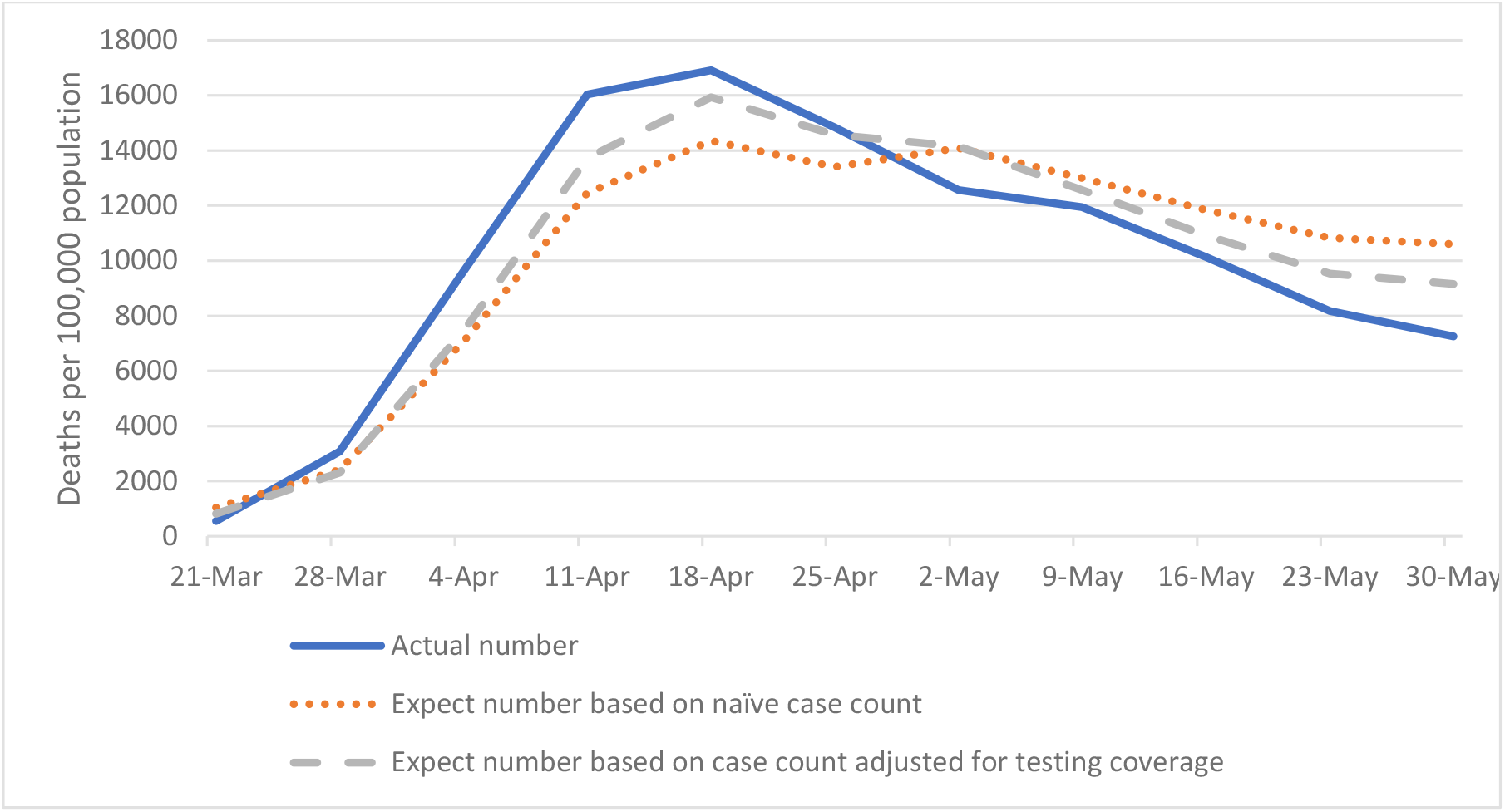
Actual and expected number of weekly US COVID-19 deaths. Notes: Linear regression was used to generate expected values of deaths based on viral testing results from the prior week. Regression results are available in appendix table 2.

## Discussion

The results of viral tests for COVID-19 provide a valuable if imperfect indicator of the severity of the disease outbreak. Compared to a simple count of cases through viral testing, a case count that is adjusted for the percentage of tests that come back positive tracks better with future COVID-19 hospitalizations and with various measures of future deaths that are likely attributable (either directly or indirectly) to the COVID-19 pandemic^17,18,19^ While some of these outcome measures may themselves be vulnerable to underreporting, estimates of excess all-cause deaths should not be affected by under-detection of COVID-19 cases, although they may be affected by behavioral changes (such as avoiding emergency room visits) that do not track perfectly with outbreak severity. The fact that this excess death measure tracks well with the prior week’s case counts adjusted for testing coverage in the manner described here supports the case for the validity of this coverage-adjusted case indicator.

Based on the estimated regression results, one might employ the following rule of thumb when comparing cases over time or across jurisdictions: for every percentage point that the positive test rate is above 0%, each newly confirmed COVID-19 case should be counted as an extra .01-.02 cases. For example, with a positive test rate of 20%, each confirmed case should count as 1.2-1.4 cases while each case would only count for 1.1-1.2 cases at a positive test rate of 10%. It should be noted that this simple adjustment to case counts will not approximate the true number of infections in a population. Rather, what this adjustment to case counts will do is likely provide for somewhat better comparisons of relative outbreak severity when examining case counts from geographic units with different levels of testing coverage, or when charting a trajectory over time for a jurisdiction that has seen changes in the adequacy of its testing coverage.

When considering the generalizability of this study’s findings, it is important to bear in mind that since the data used in this analysis was collected during a time when most areas of the United States were experiencing community transmission of COVID-19, the association between test results data and other indicators of outbreak severity may look different in the context of an effective test-and-trace system in which community spread is limited and most individuals being tested have been linked to confirmed COVID-19 cases.

Viral test results are one of the few leading indicators available to policy makers and public health officials who must respond to current local conditions based on whatever information is available, but viral testing data is still subject to substantial reporting delays which are important to bear in mind whenever interpreting indicators based on such data. While the exact length of this lag probably varies from state to state and is difficult to precisely estimate, the CDC reports that 75% of the time, the lag between symptom onset and reporting of a confirmed case to the CDC is 15 days or less.^20^ Combined with the several days it typically takes after initial infection for a patient to begin experiencing symptoms, it is not unreasonable to assume that confirmed case reports can easily lag two to three weeks behind initial infections. This timeline may be somewhat shorter if states are reporting case counts to the public prior to sending tallies of these same cases to the CDC. To account for day-of-week effects on the processing/reporting of test results as well as other noisiness from day-to-day fluctuations in case counts, case counts are often reported using a 7-day rolling average, which will effectively introduce an additional lag of a few days in the case data. Deaths and hospitalizations are subject to many of these same reporting delays and are also outcomes that typically occur later on in the progression of the disease. As such, hospitalization and death data are expected to lag even further behind confirmed case counts.

Given that viral test results become available days or weeks before high-quality mortality data is available, it is important to ensure the availability of accurate and up-to-date data on both positive and negative viral test results. A good first step toward this end would be to create better national standards in the US for reporting viral test results, including a requirement to report antibody test results separately from viral test results. Other methods of disease surveillance could also be implemented, such as conducting viral and antibody tests on random samples of the population. Test result obtained from random samples would not need to be adjusted for testing coverage and would also allow for potential detection of some presymptomatic cases, enabling earlier detection of recent infections (thereby reducing the lag time between infection and confirmed case counts). In the absence of such a testing regime, the method of adjusting confirmed case counts for testing coverage that was introduced in this study can provide a reasonable means of comparing relative prevalence based on currently-available data.

## Data Availability

Data and replication code is publicly available.

https://github.com/favero-nate/covid-underreporting

## Appendix

The case multiplier can be easily derived from the regression coefficients displayed in Table 1. The linear regression line is ŷ = β_1_c + β_2_cp (c = newly confirmed cases; p = percent positive among new tests). This can be rewritten as: ŷ = β_1_c(1 + (β_2_/β_1_)p). The adjusted case count is simply the case count times the case multiplier (cm), with the case multiplier taking the form m = 1 + αp. Thus, if α is set equal to β_2_/β_1_, then the regression line expresses the predicted value of the outcome (ŷ) as a linear function of the adjusted case count (with a y-intercept of 0).

**Appendix table 1.** Descriptive statistics.

|  | count | mean | sd | min | max |
| --- | --- | --- | --- | --- | --- |
| Newly confirmed cases<sup>a</sup> | 609 | 43.97 | 50.04 | 0.00 | 341.59 |
| Percent positive (among new tests) | 609 | 11.36 | 13.41 | 0.00 | 100.00 |
| COVID-19 deaths<sup>a</sup> | 503 | 2.54 | 3.96 | 0.00 | 33.79 |
| Excess select respiratory illness deaths<sup>a</sup> | 503 | 2.56 | 4.17 | -1.00 | 35.40 |
| Excess all-cause deaths<sup>a</sup> | 503 | 3.93 | 6.07 | -23.68 | 49.31 |
| Current COVID-19 hospitalizations<sup>a</sup> | 390 | 14.27 | 15.25 | 0.09 | 95.45 |

Notes: <sup>a</sup>indicates counts measured per 100,000 population. sd = standard deviation. min = minimum. max = maximum.

**Appendix table 2.** Ordinary least squares regression results.

|  | COVID-19 deaths |  |
| --- | --- | --- |
|  | b/se | b/se |
| Naïve number of new cases | 0.064***<br>(0.011) |  |
| Number of new cases adjusted for testing coverage |  | 0.050***<br>(0.0049) |
| Constant | 668<br>(1733) | 392<br>(1046) |
| R-sqr | 0.800 | 0.922 |
| N | 11 | 11 |

Notes: Standard errors (se) in parentheses. Both independent variables were lagged 1 week. The sample consists of weekly data from March 15-May 30, 2020. National totals for COVID-19 deaths were reported by the NCHS, and national data on new cases and testing coverage were created by aggregating data from all 50 states plus the District of Columbia and Puerto Rico. The number of new cases adjusted for testing coverage variable was computed as: $\text{number\_of\_new\_cases} (1 + 0.021 \times \text{percent\_positive\_tests})$ . The multiplier of .021 was derived from the coefficient estimates for the second model of COVID-19 deaths shown in Table 1. \* $p < 0.05$ , \*\* $p < 0.01$ , \*\*\* $p < 0.001$ (two-tailed).

